# Efficient Tracing of the SARS-CoV-2 Omicron Variants in Santa Barbara County Using a Rapid Quantitative Reverse Transcription PCR Assay

**DOI:** 10.1101/2022.07.12.22277554

**Authors:** Zach Aralis, Stewart Comer, Henning Ansorg, Carl Palmer, Jennifer Smith, Stu Feinstein, Lynn N. Fitzgibbons, Carolina Arias

**Affiliations:** Department of Molecular, Cellular, and Developmental Biology, University of California, Santa Barbara; Department of Biomolecular Science and Engineering, University of California, Santa Barbara; Santa Barbara County, Public Health Department; Department of Pathology, Santa Barbara Cottage Hospital; Pacific Diagnostic Laboratories, Santa Barbara; LegacyWorks Group, Santa Barbara; California NanoSystems Institute, University of California, Santa Barbara; Neuroscience Research Institute, University of California, Santa Barbara; Department of Medical Education and Division of Infectious Diseases, Santa Barbara Cottage Hospital; Center for Stem Cell Biology and Engineering, University of California, Santa Barbara

## Abstract

The recent emergence of the SARS-CoV-2 Omicron variant is associated with a dramatic surge of cases around the globe in late 2021 and early 2022. The numerous mutations in this variant, particularly in the Spike protein, enhance its transmission, increase immune evasion, and limit treatment with monoclonal antibodies. Identifying a community’s introduction to a novel SARS-CoV-2 variant with new clinical features related to treatment options and infection control needs is imperative to inform decisions by clinicians and public health officials, and traditional sequencing techniques often take weeks to result. Here, we describe a quantitative reverse transcription PCR assay (RT-qPCR) to accurately and precisely detect the presence of the Omicron sublineages BA.1/BA1.1 and BA.2 viral RNA from patient samples in less than four hours. The assay uses primers targeting the BA.1/BA1.1 unique mutations N211del, L212I, and L214 insertion EPE in the Spike protein gene, and the BA.2 specific mutations T19I and L24/P25/P26 deletion in the Spike protein gene. Using this assay, we detected 169 cases of Omicron, 164 BA.1/BA1.1 and 5 BA.2, from 270 residual SARS-CoV-2 positive samples collected for diagnostic purposes from Santa Barbara County (SBC) between December 2021 to February 2022. The RT-qPCR results show concordance with whole viral genome sequencing. Our observations indicate that Omicron was the dominant variant in SB County and is likely responsible for the surge of cases in the area during the sampling period. Using this inexpensive and accurate test, the rapid detection of Omicron in patient samples allowed clinicians to modify treatment strategies and public health officers to enhance contact tracing strategies. This RT-qPCR assay offers an alternative to current variant-specific detection approaches, provides a template for the fast design of similar assays, and allows the rapid, accurate, and inexpensive detection of Omicron variants in patient samples. It can also be readily adapted to new variants as they emerge in the future.

## Introduction

SARS-CoV-2, the causative agent of the COVID-19 pandemic, is a single strand positive RNA virus of the coronavirus family. COVID-19 has caused a devastating number of cases and deaths since it was officially declared a pandemic on March 11th, 2020. Despite the development of successful vaccines and global vaccination efforts, SARS-CoV-2 continues to spread globally^1–3^. A challenge in controlling COVID-19 is the emergence of SARS-CoV-2 variants as mutations accumulate in the viral genome. While many of these mutations are of little to no consequence, others can provide a higher viral fitness by increasing virus transmission efficiency, conferring resistance to immune response, and/or impacting the severity of the disease^4–6^. SARS-CoV-2 variants that pose a significantly increased risk to the global community are designated Variants of Concern (VOC) by the World Health Organization (WHO)^7^. The control of SARS-CoV-2 transmission relies on our capacity to diagnose COVID-19 efficiently, detect the emergence of new viral lineages, and determine variant prevalence in the population^8–11^.

One of the most aggressive SARS-CoV-2 variants identified, Omicron BA.1 (originally B.1.1.529), was initially detected in South Africa in November of 2021, where it rapidly outcompeted other viral variants in the region^12^. In November 2021, the WHO classified SARS-CoV-2 Omicron as a VOC and linked it to a global upsurge of cases at the end of 2021^13^. By December 2021, the highly transmissible Omicron variant caused more than half of all daily SARS-CoV-2 infections^14^. Omicron carries numerous mutations, including 30 in the Spike (S) protein, enhancing binding to the receptor ACE2 and increasing immune evasion^15–20^. The BA.1.1 sublineage has all mutations of BA.1 and as well as the R346K substitution^21^.

With the high transmission rates of Omicron, it was anticipated other sublineages of this variant would emerge. Of public health relevance, the Omicron sublineage BA.2, first detected in December 2022, is a highly contagious variant classified as a VOC in February 2022^22^. While the global prevalence of BA.2 was lower than that of BA.1/BA.1.1 through January and February of 2022, it accounted for over 50% of the global cases sequenced by the first week of March. Several mutations in the S protein distinguish BA.2 from BA.1/BA.1.1 and may enhance the transmissibility and immune evasion of this variant^22^.

The numerous mutations in the S protein in BA.1/BA1.1 and BA.2 limit the options for treatment; of the three monoclonal antibody treatments approved for early use by the FDA, only Sotrovimab is effective against the BA.1/BA.1.1 Omicron variant. However, this monoclonal antibody is ineffective in treating the BA.2 Omicron Variant^23^. While testing each patient to identify the infecting viral variant and tailor individual treatment is an unviable option, understanding the prevalence of specific variants in the community can inform decisions about using monoclonal antibodies and other COVID-19 therapies.

Next-generation sequencing (NGS) of the viral genome is the standard method to determine the SARS-CoV-2 variant present in a sample. While NGS provides a wealth of information on the mutations present in an individual strain of the virus, it typically takes days to weeks to accurately identify viral variants. Rapid, simple, and high throughput methods to identify Omicron and other emerging viral variants are urgently needed to provide clinicians and public health officers with essential real-time information on the prevalence of specific SARS-CoV-2 variants in the population.

Here we developed an RT-qPCR assay to distinguish between the Omicron BA.1/BA.1.1 and BA.2 variants of SARS-CoV-2 by targeting a set of specific mutations in the S1 domain unique to each of them. Using this assay, we obtained one hundred percent concordance with NGS for Omicron variant identification, and we accurately detected the dominance of the Omicron variant in Santa Barbara County. Active communication with our local public health department and hospitals and the immediate dissemination of these data with health officials and clinicians provided vital information to treat active cases in the region and help manage the dramatic rise in COVID-19 cases in Santa Barbara County.

## Methods

### Primer Design

Primers were designed based on the SARS-CoV-2 Wuhan-Hu-1 (Wu-Hu1) genome (accession number NC_044512.2). For BA.1/BA.1.1 specific targets, the sequence was modified to include a 3-nucleotide deletion at position 22194-22196, and a 9-nucleotide insertion previously reported for this variant^24,25^. For BA.2, specific primers were designed to target a 9-nucleotide deletion at position 21633–21641 and the lack of a 6-nucleotide deletion at amino acids 69/70 present in BA.1/BA.1.1 and other VOCs^26^. The positions of the primers are illustrated in Figure 1 and the primer sequences are shown in Table 1.

**Table 1:**
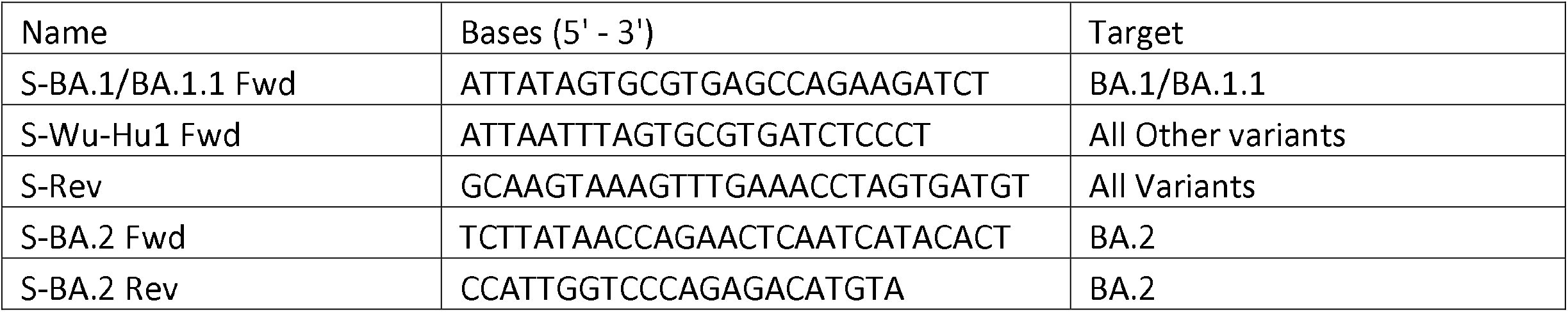
Primer sequences used in this study

**Figure 1.**
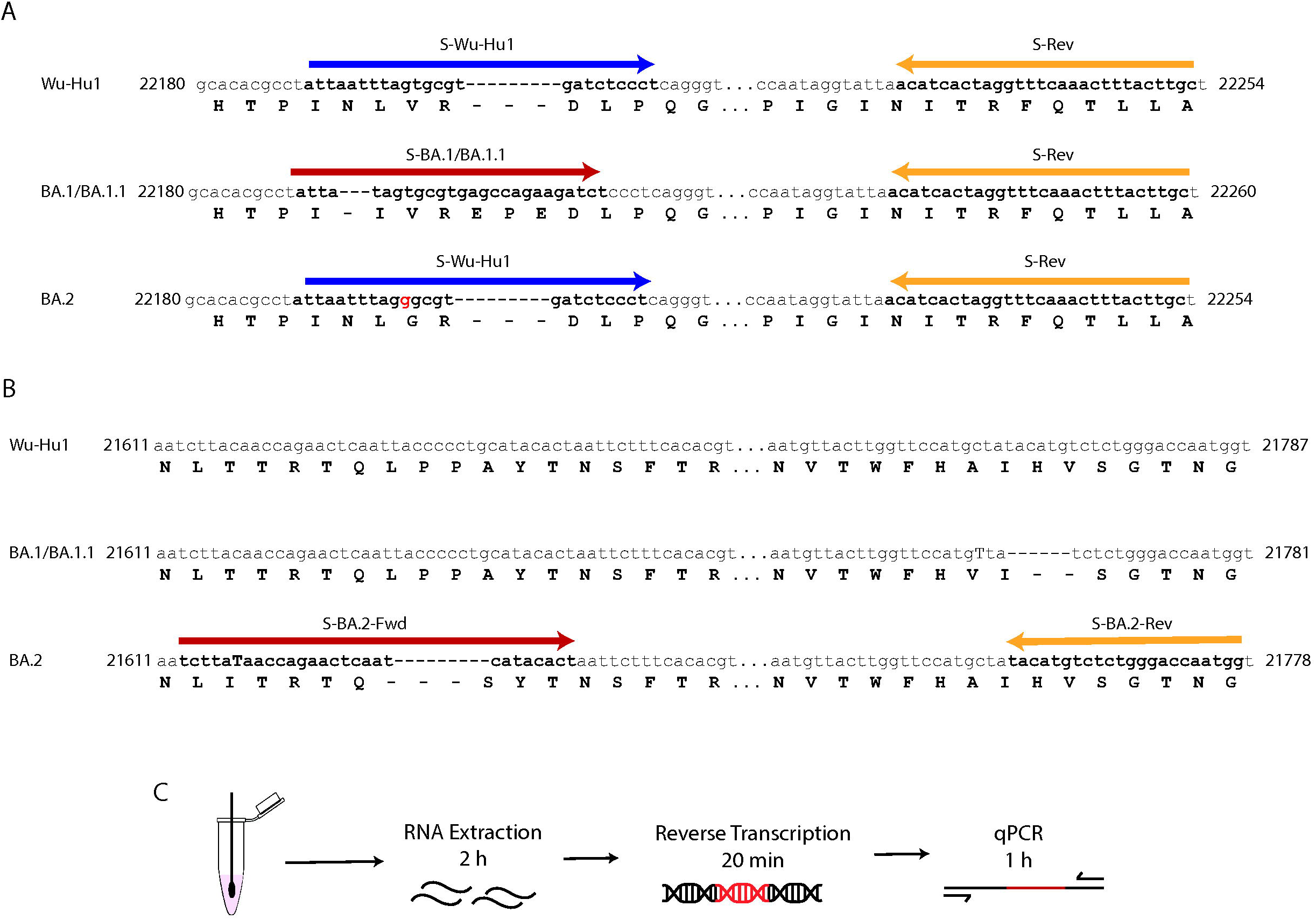
Primer Positions and Method overview. Genomic coordinates of the primer binding locations in the Wu-Hu1, BA.1/BA.1.1, and BA.2 viral variant genomes for (A) the BA.1/BA.1.1 targets and (B) the BA.2 targets. (C) Schematic and estimated timeline of RT-qPCR assay.

### Sample collection and RNA extraction

Clinical samples were acquired as residual NP swabs stored in Universal Transport Media (UTM) or Viral Transport Media (VTM). Samples were inactivated at 56 °C for 30 minutes, and RNA was extracted using the QIAamp MinElute Virus Spin Kit [Qiagen, 57704] from 140µL of the sample and eluted in 50 µL. The Santa Barbara Cottage Hospital IRB reviewed and approved pre-and post-analytical protocols. This study followed the Strengthening the Reporting of Observational Studies in Epidemiology (STROBE) reporting guideline for cohort studies.

### RT-qPCR

Viral RNA was reverse transcribed by mixing 8 µL of extracted RNA with 2 µL of LunaScript RT SuperMix [NEB, E3010L], followed by incubation using the thermal profile: 25°C for 2 minutes, 55°C for 10 minutes, and 95°C for 2 minutes. A qPCR master mix was prepared by combining 5 µl of nuclease-free water, 2 µl of 5µM S-Omicron or S-Wu-Hu1 Fwd primer, 2 µl of 5µM S-reverse primer, 10 µl of the PowerUp™ SYBR™ Green Master Mix [Applied Biosystems, A25741], and 1 µL of cDNA, for a total reaction volume of 20 µL. All components were gently mixed by pipetting, and the reactions were collected by centrifugation using a tabletop centrifuge. The reactions were run on a Bio-Rad CFX96 Touch using the following thermal protocol: 50°C for 2 minutes; 95°C for 2 minutes; 40 cycles of 95°C for 15 seconds followed by 60°C for 1 minute, and the plate read in the SYBR/Fam channel. Data were analyzed using the Bio-Rad CFX Maestro software with a single threshold to determine the quantification cycle.

### Data Interpretation

The assay was valid if samples had a Ct value of 37 or below in the reactions with the S-BA.1/BA.1.1, BA.2 or S-Wu-Hu1 reaction. This cutoff is the Ct value below by which all samples could be successfully determined via NGS. Samples were defined as SARS-CoV-2 BA.1/BA.1.1 or BA.2 if the Ct value for the sample was equal to or lower than 37 in the qPCR reaction, including the S-Omicron BA.1/BA.1.1 or BA.2 Fwd primer. In the qPCR reaction, samples with Ct values above 37, including the S-Omicron or S-Wt primers, were called inconclusive. A small number of samples with low Cts for one target had background amplification for one of the other targets. In these cases, if the difference between Ct values was greater than 10 Ct, then the target with the higher value was called as not determined. This was likely caused by non-specific amplification typically observed in mutation-specific qPCR assays^27–29^.

### Amplicon Library Generation, Next Generation Sequencing, Phylogenetic Analysis

Viral cDNA was generated using LunaScript RT SuperMix, followed by incubation using the thermal profile: 25°C for 2 minutes, 55°C for 10 minutes, and 95°C for 2 minutes. The cDNA was amplified using the SARS-CoV-2 genome tiling primer pools from the UCSF CAT COVID-19 Tailed 275bp ARTIC V3 Protocol.^30^ The PCR reaction was prepared by mixing 5µL of Q5 Hotstart 2X Master Mix [NEB, M0494S], 1.8µL of primer pool 1 or 2, 2.2µL of nuclease-free water, and 1µL of cDNA per sample. Amplification was carried out at 98°C for 30 seconds, followed by 35 cycles of 95°C for15 seconds and 63°C for 5 minutes. Pools 1 and 2 were combined, diluted 1:100 in nuclease-free water, and indexed using NEBNext dual index oligos for the Illumina [NEB, E6440S]. Samples were equal volume pooled, cleaned up with AMPureXP beads, and sequenced on a NextSeq 500 using a NextSeq 500/550 Mid Output Kit v2.5 (300 Cycles) kit. The demultiplexed FASTA files were uploaded to CZ ID for alignment. Consensus sequences were uploaded to Pangolin COVID-19 Lineage Assigner for variant identification. Consensus sequences generated by NGS were uploaded to Nexstrain for phylogenetic analysis^31^. Phylogenetic trees were visualized with the ggtree package in R^32^.

## Results

The Omicron variant of SARS-CoV-2, initially reported at the end of November 2021 in South Africa, is characterized by numerous mutations throughout the genome. Most of these mutations, 30 in BA.1/BA.1.1 and 20 in BA.2, have been described in the Spike (S) protein gene. To develop a quantitative reverse transcription PCR (RT-qPCR) assay to precisely detect the Omicron variant sublineages BA.1/BA.1.1 and BA.2 in clinical samples, we designed primers targeting unique mutations present in the S gene of each of these variants. The BA.1/BA.1.1 primers target the N211 deletion (N211del), the L212I substitution, and the 214 EPE insertion (ins214EPE) (Fig 1A). The BA.2 primers target the T19I substitution and L24/P25/P26 deletion (Fig 1B). In parallel, we designed the S-Wu-Hu1 primer targeting the region in the S gene encoding for amino acids (AA) 210-217 that would recognize all SARS-CoV-2 variants of concern, except for Omicron BA.1/BA.1.1 (Fig 1A). Our RT-qPCR assay takes less than 4 hours from RNA extraction to readout (Fig 1C). Notably, we successfully designed and validated the assay and obtained results from patient samples within one week (Fig 2A).

**Figure 2.**
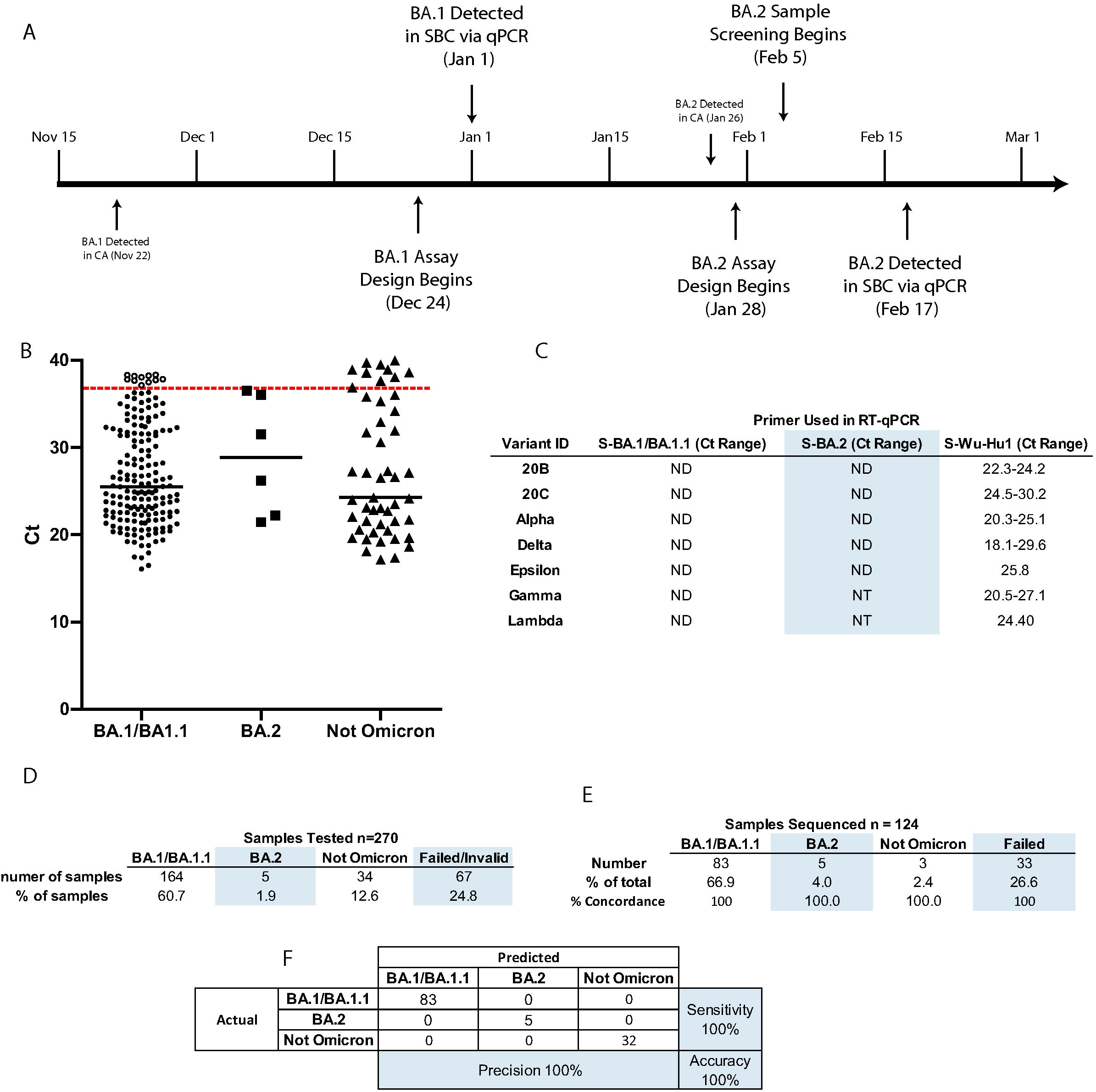
Design timeline and metrics of the Omicron variant-specific RT-qPCR. (A) Timeline of events around the design, optimization, and implementation of the Omicron variant-specific RT-qPCR in Santa Barbara County. (B) Ct values for all samples tested. The red dashed line indicates the cutoff at a Ct of 37. (C) Ct values of a panel of samples confirmed to be variants other than Omicron BA.1, BA.1.1, and BA.2. (D) Results of all 270 samples tested. (E) Percent concordance and (F) confusion matrix of samples with whole viral NGS and variant-specific RT-qPCR data. ND: not detected; NT: not tested.

To validate this assay, we obtained 270 residual SARS-CoV-2 positive nasopharyngeal swab samples collected for diagnostic purposes between December 2021 and February 2022, with undetermined viral variant (Fig 2A. Timeline). As a control, we used 29 retrospective residual SARS-CoV-2 positive nasopharyngeal swab samples we previously identified by viral genome sequencing as 20B, 20C, Epsilon (CAL.20C), Gamma (P.1), Lambda (C.37), Alpha (B.1.1.7), and Delta (B.1.617.2 and AY) (Fig 2B). Using our RT-qPCR assay in this cohort, we detected BA.1/BA.1.1 in 164 samples (60.7% of total), BA.2 in 5 samples (1.9% of total), and other variants in 34 samples (12.6% of total). Our results confirmed the overwhelming presence of SARS-CoV-2 Omicron in most samples collected in late 2021/early 2022. The 67 remaining samples failed due to a lack of amplification and detection by RT-qPCR with the S-Omicron or the S-Wu-Hu1 primers, possibly due to low viral load or poor sample preservation (Fig 2D). The 20B, 20C, Alpha, Delta, Epsilon, Gamma, and Lambda control samples were detected by RT-qPCR with the S-Wu-Hu1 but not with the S-Omicron BA.1/BA.1.1 specific primer (Fig 2C).

To confirm the presence of the Omicron variant in these clinical specimens and evaluate the accuracy of our assay, we sequenced the viral genome in 124 of the 270 patient samples that were tested using our RT-qPCR assay. We identified 83 SARS-CoV-2 Omicron BA.1/BA.1.1 (91.2% of samples successfully sequenced), 5 Omicron BA.2 (5.5%), and 3 SARS-CoV-2 Delta (3.3%) (Fig 2E). Thirty-three samples failed viral genome sequencing, likely due to low viral load or poor sample preservation. The identification of the SARS-CoV-2 variants using whole-genome sequencing showed one hundred percent concordance with our RT-qPCR assay identifications, highlighting the specificity and accuracy of our assay (Fig 2F). Genetic and phylogenetic analyses of the sequenced genomes show defining mutations of the omicron sublineages (Fig 3) and the presence of three distinct clusters corresponding to BA.1, BA.1.1, and BA.2. The sublineages BA.1 and BA.1.1 were introduced late in 2021 and continued circulating in the Santa Barbara County (SBC) population throughout early 2022. We detect the introduction of BA.2 in the 6^th^ week of 2022, with sustained transmission until the end of our sampling period (Fig 4A, 4C). Our phylogenetic analyses support the local transmission of these variants.

**Figure 3.**
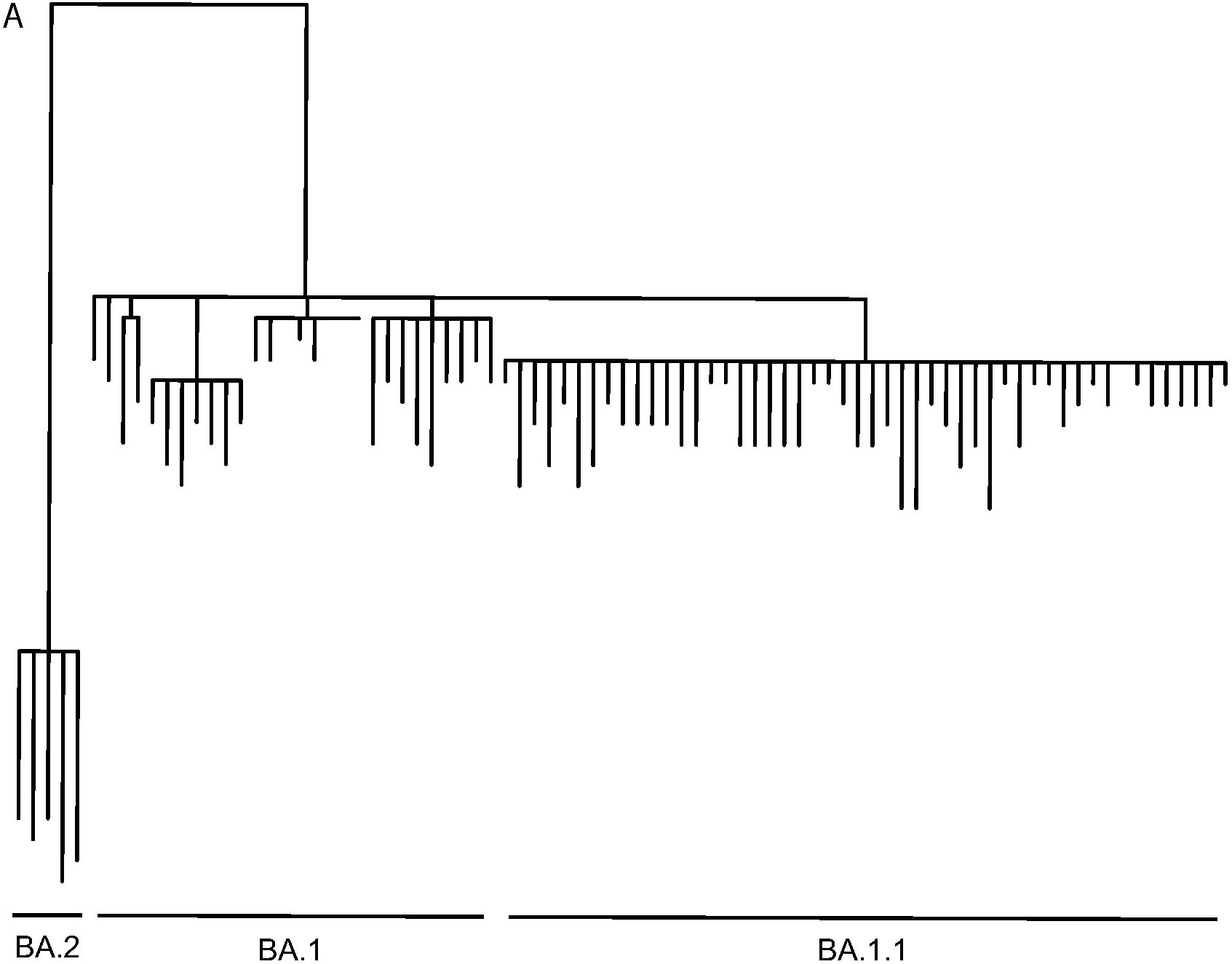
Phylogenetic analysis based on whole-genome sequences of SARS-CoV-2 Omicron variants detected in Santa Barbara County.

**Figure 4.**
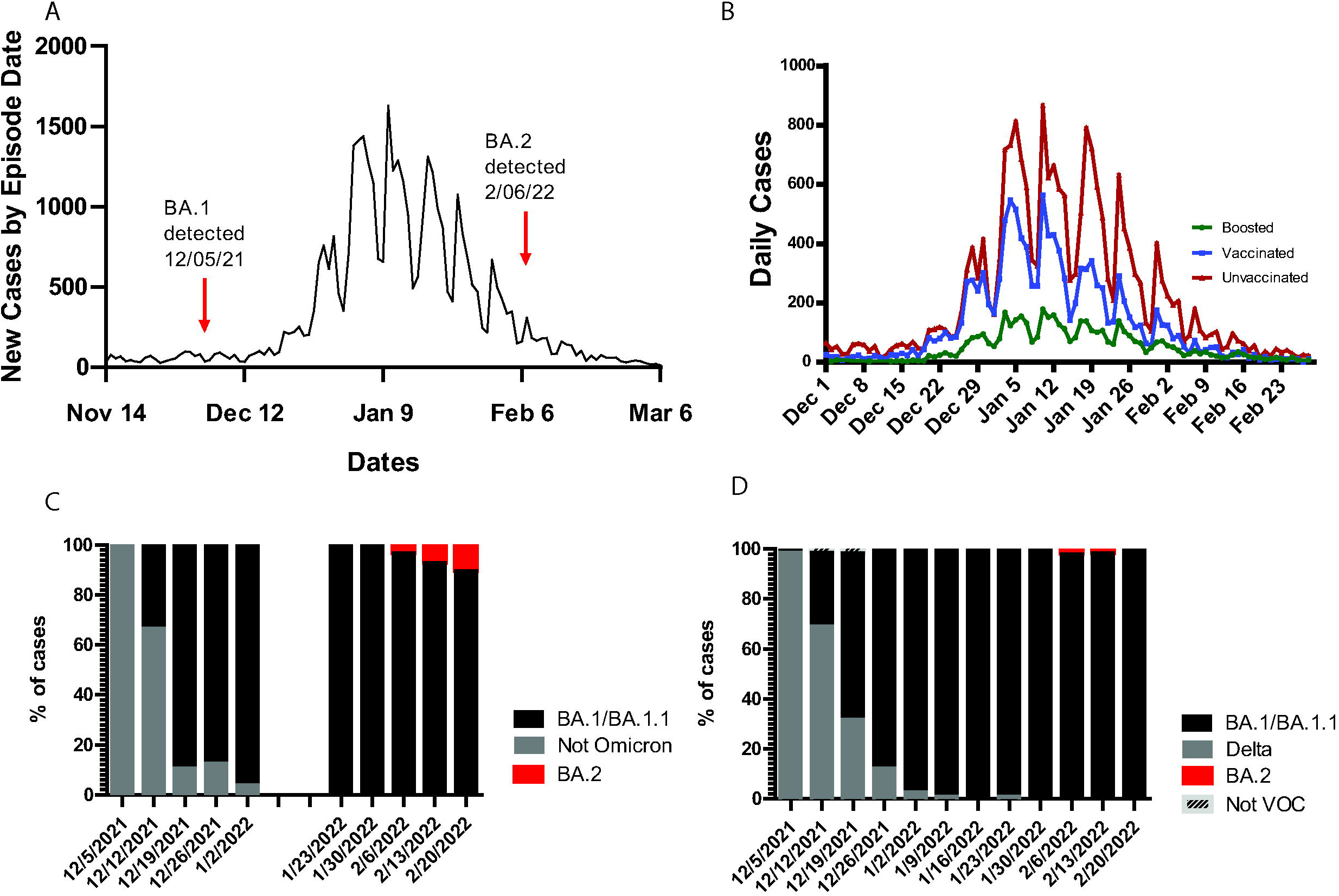
Daily cases and relative variant proportions in Santa Barbara County. (A) Total new COVID-19 cases by episode date and (B) new cases in boosted, vaccinated, and unvaccinated individuals in Santa Barbara County reported by the SBCPHD^34^. (C) Relative proportion of omicron samples in Santa Barbara County, determined via variant specific RT-qPCR and (D) whole viral NGS.

The presence of Omicron (BA.1, BA.1.1, and BA.2) in the clinical samples from SBC is linked to a dramatic surge in the number of cases since December of 2021 mainly in the unvaccinated population (Fig 4A, 4B). The weekly distribution of SARS-CoV-2 variants in clinical samples, determined by qRT-PCR (Fig 4C) or whole genome sequencing (Fig 4D), shows the dominance of the Delta variant in SBC throughout November and the initial detection of Omicron in the week of December 5th, 2021. The rapid expansion of the Omicron variant in the population is seen in the following two weeks, with the complete replacement of the Delta variant and the dominance of Omicron present in 100% of the samples tested by the first week of January 2022 (Fig 4C). We detected the BA.2 sublineage during the second week of February 2022, with low prevalence (below 5%) throughout the rest of the month.

## Discussion

Here we report developing a rapid, specific, and accurate RT-qPCR-based method to detect and distinguish between Omicron sublineages BA.1/BA.1.1 and BA.2 in patient samples. As new SARS-CoV-2 variants continue to emerge and fuel COVID-19 cases worldwide, it is critical to have tools for the rapid identification and characterization of viral variants at hand, to inform both clinical and public health decisions. While other assays specifically detect BA.1/BA.1.1 and BA.2, none were developed and implemented in a time frame to impact the health and safety of the local community. Similar diagnostic tests typically take weeks to months to be developed and implemented. However, our open collaboration with local health officials allowed us to develop, validate, and use this diagnostic assay in clinical samples and provide retrospective and current Omicron prevalence data in under one week. This same strategy and assay design is readily adaptable for future use as new variants emerge.

In Santa Barbara County, our early adoption of next-generation sequencing methods for genomic surveillance of SARS-CoV-2 became a central tenet of the local COVID-19 pandemic response. NGS of viral genomes shed light on the regional distribution and prevalence of viral variants and informed prevention and clinical intervention strategies^33^. While informative and critical for our response, state-wide and local next-generation surveillance initiatives provided results with a one to six weeks delay.

The rapid emergence and high transmissibility of Omicron presented the risk for a precipitous local surge as seen in other communities. The successful control of potential Omicron outbreaks required the immediate detection of this variant in patient samples. Moreover, from the clinical management perspective, knowing the presence and prevalence of Omicron in our patient population can help guide the selection and use of limited-supply monoclonal antibody therapies. Considering the urgent need to detect the Omicron variant in our community, we collaborated with SBC public health officers and clinicians to develop and validate a rapid assay to differentiate the BA.1/BA.1.1 Omicron sublineage from other SARS-CoV-2 variants present in patient samples.

In this assay, based on a simple RT-qPCR reaction, we took advantage of the unique mutations present in BA.1/BA.1.1 and designed primers spanning the deletion in position 211 (N211del) and the insertion in position 214 (L214 EPE). Targeting these short but significant changes in the viral genome ensures that PCR amplification will only occur when the primer binds the cDNA derived from the BA.1/BA.1.1 genome, and no other variant. This rapid assay (4 hours from start to completion) requires basic equipment and reagents commonly found in molecular biology laboratories, is scalable, and demands minimal optimization. Notably, parallel viral genome sequencing of patient samples tested by RT-qPCR confirmed the accuracy and specificity of the assay for BA.1/BA.1.1 detection.

Using the RT-qPCR assay to test samples from December 2021 to January 2022, we showed the potential introduction of Omicron BA.1/BA.1.1 in the SBC community during the first week of December. The rapid transmission of BA.1/BA.1.1 was reflected in its close to 90% prevalence and its dominance of the variant landscape in our community two weeks after it was first detected. These valuable data informed clinicians and discouraged the use of monoclonal antibodies to manage active cases. Moreover, identifying Omicron in the SB population triggered enhanced testing and contact tracing to manage viral transmission.

In December 2021, as the cases of COVID-19 attributed to BA.1/BA.1.1 continued to rise globally, variant surveillance studies reported the emergence of a new Omicron sublineage known as BA.2. This variant differs from BA.1/BA.1.1 by approximately 27 mutations, is more transmissible, and is considered a VOC. Due to the potential risk of continued viral transmission and new case surges fueled by BA.2, we rapidly adapted our RT-qPCR assay to detect this variant specifically, reducing the turnaround time from weeks for sequencing to hours for this targeted PCR. Following our initial design for the BA.1/BA.1.1 assay, we modified our primers to recognize the unique deletion in AA positions 24/25/26 present in BA.2. As with our initial iteration, the design, validation, and initial tests of patient samples spanned one week (01/28/22 to 02/05/22). Our surveillance efforts identified the first case of BA.2 in SBC on February 17th, 2022, in samples collected for COVID-19 diagnosis the week before variant testing. We immediately reported these results to clinicians and public health officials, who used this information to enhance contact tracing and monitor the first outbreak of BA.2-associated cases in SBC. The identity of the BA.2 variant in the samples tested was confirmed by whole viral genome sequencing, validating the accuracy and specificity of our assay to detect this Omicron sublineage. The success of the detection of BA.2 highlights the straightforward design and adaptability of this assay and provides a template for the fast identification of emerging SARS-CoV-2 variants in patient samples.

As the COVID-19 pandemic continues, rapidly deployable variant detection methods will remain indispensable for enhancing public health. Our results demonstrate that a simple variant assay, needing minimal troubleshooting and optimization, successfully captures the introduction of SARS-CoV-2 variants in the population and provides valuable real-time data during emerging surges to guide clinical and public health efforts. Most importantly, this study illustrates the power and impact of collaborative work and open communication between academic research laboratories, clinicians at local hospitals, and public health officers to rapidly respond to a public health emergency, significantly improving outbreak control responses and public safety.

## Data Availability

All data produced in the present study are available upon reasonable request to the authors.

## Acknowledgments

The authors acknowledge the contributions of the Biological Nanostructures Laboratory within the California NanoSystems Institute at the University of California, Santa Barbara. We also thank the University of California Santa Barbara, the University of California, Office of the President, the CDC’s COVID-NET initiative and the Cottage Health Research Institute for financial support. We thank Osvaldo Moreno, Adriana Ramirez Negron, Dylan David, and Julia Conrad for technical support. We thank Joy Kane and the entire Santa Barbara County epidemiology team for insightful discussions. Lastly, we thank clinicians and laboratory staff at Marian Hospital, Sansum Clinic, UCSB Student Health, Cottage Hospital, and the Santa Barbara Department of Public Health for providing samples, critical discussions, and support for this project.

